# A Feasibility Trial to Evaluate the Composite Efficacy of Inhaled Nitric Oxide in the Treatment of Covid 19 Pneumonia : Impact on Viral Load and Clinical Outcomes

**DOI:** 10.1101/2021.04.15.21255300

**Authors:** Merlin Moni, Thushara Madathil, Dipu T Sathyapalan, Veena Menon, Georg Gutjahr, Fabia Edathadathil, Deepthi Sureshkumar, Preetha Prasanna, Soumya Jose, Roshni Jerome, Ajai Krishnan, Indulekha C Pillai, Geetha Kumar, Bipin Nair, Aveek Jayant

## Abstract

**Background:** Hypoxic patients with Covid 19 pneumonia are at high risk of adverse outcomes. Inhaled Nitric Oxide (iNO) inhibits viral entry and replication of SARS-CoV2 and *in vivo* proof of its antiviral actions is unavailable to date. This feasibility study was conducted to test the antiviral effects of iNO and to describe clinical outcomes.

**Trial design and Methods:** The phase II open label, randomised controlled feasibility trial(ISRCTN 16806663) conducted at a South Indian tertiary care referral centre, recruited COVID-19 pneumonia patients with hypoxic respiratory failure and allocated them into iNO cases and control groups(1:1). iNO was administered as pulses for 30 minutes for three consecutive days at 12-hour intervals in cases, in addition to standard of care received by the control group. The primary outcome was decline in viral load, as defined by a surrogate change in the RT-PCR cycle threshold. The co-primary clinical outcome was time to improvement of >2 points on the WHO Ordinal Scale(WOS).

**Results:** Among the 29 patients enrolled, 14 iNO cases and 11 controls completed the study protocol. Longitudinal analysis revealed a significant difference in the decline (p <0.002, N= 23) in viral load among the iNO cases compared to controls. The proportion of patients achieving 2-point improvement in the WOS within 14 days of randomisation was significantly higher in the iNO cases (n=11, 79%), as compared to the controls (n=4, 36%) (p=0.05).

**Conclusions:** Our study demonstrated significant improvement in virological and clinical outcomes among patients with adjunct iNO therapy and no adverse effects were reported.

## BACKGROUND

The current SARS CoV-2 pandemic has been a health emergency of international concern for over an year (1). The virus has caused over 100 million infections and more than 2 million deaths and the pandemic continues to smoulder. Given the pandemic is still evolving with the advent of multiple variants and consequent waves of outbreak reported across the globe, there is an existing need for effective therapies. In pandemics, therapeutics typically turn to repurposing established medications(2). Efforts to repurpose drugs as anti- virals in Covid -19 have not yielded spectacular results(3). The maximum benefit has actually been derived dexamethasone which modulates host immune response to the virus(4). However, viral loads are of clinical relevance as high viral loads and their persistence is linked to severity of disease (5) and the efficacy of an anti-viral is often best when initiated in the early phase of the disease.. This concept has been validated in earlier models of virus-induced Severe Acute Respiratory Infections (6) and secondary analyses of the Adaptive Covid-19 Treatment Trial (7)(8)(9)

Nitric oxide (NO) is a biologic molecule with pleotropic effects and wide presence across mammalian organ systems. (10). It performs a key role in maintaining vascular homeostasis(10)(11). NO or its donors have been used as anti-anginals (12) and in pulmonary hypertension (13). NO also possesses potent oxidant potential that is linked to putative harm (14) but also makes it a potent broad spectrum antimicrobial (15). NO donors have been shown to previously inhibit viral protein and ribonucleic acid synthesis in a model of Coronavirus causing Severe Acute Respiratory Infection (16), and, more recently, in SARS CoV2 as well (17). NO has also been shown to reduce palmitoylation of the nascent spike (S) protein, which affects the fusion between this protein and its cognate receptor, angiotensin converting enzyme-2 (ACE-2), which is the primary step mediating viral entry into host cells (18). There has also been suggestive clinical evidence of its utility in SARS caused by Coronaviridae earlier (6). It is unsurprising that there have been efforts to repurpose this molecule for SARS CoV-2 (15)(19).

Inhaled NO has actually been the subject of investigation in several reports of therapies in Covid 19. However, the overwhelming focus has been on the hemodynamic, gas exchange or endothelial modulation effects of this drug (20)(21)(22)(23)(24)(25), rather than as an antiviral(26). We therefore designed a randomised controlled trial to test the anti- viral effects of inhaled nitric oxide. Regulatory permission was provided only for a limited feasibility trial. We report below the results of this feasibility trial(ISRCTN16806663).

## METHODS

### Design

The study was conducted as a Phase II open-label, randomised controlled feasibility trial at a single tertiary referral centre in South India. Consent was obtained from the Institutional Ethics Committee of the Amrita Institute of Medical Sciences and Research Centre, Kochi and the Drug Controller General of India for permission to repurpose iNO. A description of the trial is also found on the ISRCTN registry (ISRCTN no 16806663).

### Participants

All consenting adults (18-60 years of age) who presented to our centre with a confirmed diagnosis of SARS CoV-2 infection (using one or more of the following tests: antigen testing (SD Biosensor, South Korea), Polymerase Chain Reaction (PCR) for SARS CoV-2 (details below) or the GeneXPert molecular platform (Xpert, Cepheid CA) AND moderate respiratory symptoms, deemed to be secondary to Covid 19. For assignment to this category of severity of disease patients needed to demonstrate room air saturation on pulse oximetry ⍰ 94% and a respiratory rate > 24 breaths/min (27)(28). Patients were excluded if they had mental obtundation and/or other contraindications to non invasive ventilation (NIV), unwillingness to receive NIV, were deemed candidates for invasive mechanical ventilation at the time of screening, Kidney Disease Improving Global Outcomes (KDIGO), Stage II or higher renal failure (29), a prevailing diagnosis of chronic renal failure, known Glucose 6 Phosphate dehydrogenase deficiency or a baseline methemoglobin >3%, hemoglobinopathies, mean arterial pressure <65 mmHg, presence of baseline pulmonary artery hypertension (as adjudged by a tricuspid regurgitation velocity of >2.8 m/s on resting trans thoracic echocardiography), or if they were pregnant or lactating.

### Randomisation and masking

Randomisation was performed by a computerised random number generator, and allocation was concealed in sealed opaque paper envelopes. While the attending medical team recruited patients, obtained consent and documented the clinical data, a separate trial team administered the inhaled nitric oxide and trial allocation envelopes were opened by the latter. The laboratory obtaining and analysing specimens was blinded to allocation. However no placebo gas was used and allocation to trial or control was visible both to the trial and attending teams.

### Procedures

All enrolled patients received care in a designated intensive care unit (ICU), where standard variables such as electrocardiography, non-invasive and/or invasive blood pressure and pulse oximetry, were continuously measured and recorded in a nursing chart. Additional monitoring was instituted according to the discretion of the ICU physician: the attending intensivist and care physicians decided on adjunct therapies such as fluid, vasopressors, non-Covid 19 related medications (if any, including anti-bacterials) and prescribed laboratory testing as they deemed fit. Patients (n=29) were allocated to either treatment with inhaled nitric oxide (study group), or control. Both groups received dexamethasone in line with the data from the RECOVERY trial and the local health guidelines (27)(28) The trialist team, distinct from the ICU team, inserted invasive radial arterial catheters to obtain baseline arterial blood gases and levels of Methaemoglobin. The trial group received inhaled nitric oxide delivered through a tight fitting face mask and the V60 respiratory assist system in pulses as below for three consecutive days post enrolment. This dose was administered in a crescendo-decrescendo fashion as follows:-

10 ppm, 0-5 minutes

20 ppm, 5-7 minutes

30 ppm, 7-9 minutes

50 ppm, 9-11 minutes

80 ppm, 11-23 minutes

80 ppm-0 ppm, 23-30 minutes, decreased at 10 ppm/minute

Nitric oxide was delivered on the V60 platform using continuous positive airway pressure values of 5-10 cm H_2_O. Study patients also received oral Sildenafil at doses of 10 mg, thrice daily (to primarily prevent iNO rebound), for 5 days from trial enrolment. iNO therapy was discontinued if the baseline or individual session methaemoglobin exceeded 3%, or serially obtained mean arterial blood pressures fell below 60 mm Hg for >15 minutes, as this warranted discontinuing Sildenafil therapy. iNO was also discontinued if a subject was transitioned to invasive mechanical ventilation. Methaemoglobin values were measured at the end of each of the 6 sessions of therapy and for 24 hours post cessation of therapy as a safety measure. During and after this trial period, additional therapies such as steroids (Dexamethasone), antivirals (remdesivir and favipiravir), antibiotics, fluid restriction, non-invasive ventilation (and/or transition to invasive ventilation) and anticoagulation were also considered and prescribed on an as needed basis by the immediate care team.

Viral load assessments were performed as follows: Nasopharyngeal (NP) swab samples collected on baseline (day 0) and thereafter on days 3, 5, 7, 10 and 14 post-treatment, were measured for SARS CoV2 RNA. Briefly, viral RNA was extracted from the NP swabs using the QIAamp® viral RNA kit (Qiagen, Hilden, Germany). The PerkinElmer® SARS-CoV-2 RT-qPCR kit (CE/IVD) (PerkinElmer, Turku, Finland), which targets two specific genomic regions of SARS-CoV2, namely the nucleocapsid (N) gene and ORF1ab, was used to assess qualitative detection of viral RNA in study samples. The limit of detection (LoD) of this kit was 1 copy/µL (3 log10 copies/ml). According to the protocol of RT-qPCR kit, a Ct value of ≤ 40 for either one of the two targets was defined as ‘positive’ while a Ct > 40 was considered ‘negative’. The cycle threshold (Ct) value of the N gene or ORF1ab gene of SARSCoV-2 is considered to inversely correlate with the viral load. In this study, viral load was calculated by the comparative Ct (ΔCt) method. The Ct value of the human RNAse P gene was considered as reference (internal control), to normalize for sampling variations. For the analysis of ‘time to negativity’ between the two groups, a cut off of Ct >35 was defined as ‘absence’ of viral loads / virus particles (30).

### Outcomes

The primary outcome was a decline in viral load as defined by a surrogate change in the cycle threshold. The co-primary outcome was the time to improvement of > 2 points on a 8 point ordinal scale formulated by the WHO Research and Development Blueprint group (31).

The defined secondary outcomes were 28 day all cause mortality, need for transition to invasive ventilation, duration of non invasive ventilation (if applicable), duration of oxygen therapy, length of hospital and intensive care unit stay, a change in the Sequential Organ Failure Assessment (SOFA) score from enrolment to 72 hours thereafter, and incidence of methaemoglobin levels >3% during the trial.

Viral loads were serially assessed on nasopharyngeal specimens obtained at enrolment, and on days 3, 5, 7, 10 and 14 thereafter, or till a Ct >35 whichever was earlier.

### Statistical Methods

Baseline characteristics were summarized by means and standard deviations. T-tests were used to compare quantitative variables between groups, Fisher’s exact tests were used to compare qualitative variables and logrank tests were used to compare time-to-event data between groups. Kaplan-Meier estimates were used to estimate and visualize event times. Longitudinal mixed-effects regression models with a random effect for the individual intercept were used to estimate the additional daily improvements in the treatment group over the control group for the dependent variables, WHO ordinal scale (treating the scale as a quantitative variable) and viral loads. Restricted splines with three interior knots were used for the dependency on time. F-tests were used to test for significance of difference groups of parameters in the longitudinal models. Multiple imputation was used to deal with missing data. MANOVA was used to test for differences between groups in SO2 and PF values. A Logistic mixed-effects model was used to test for differences in hypoxemia (SO2 < 90).

## RESULTS

### Baseline characteristics

Of the total 29 patients enrolled, 25 patients completed the trial (Figure 1). 11 of these patients received standard treatment (control group), while 14 patients received iNO in addition to the standard treatments prevailing during the study enrolment. The baseline characteristics of the cohort are given in Table 1.

**Table 1:** Baseline characteristics of patients in the control and the treatment group.

| **Characteristics** | **Control**<br>**(N=11)** | **Treatment**<br>**(N=14)** | **p-value** |
| --- | --- | --- | --- |
| Age, mean (SD) | 65.9 (10.8) | 53.8 (10.1) | < 0.01 |
| Sex, % women | 5 (45%) | 2 (14%) | 0.18 |
| SOFA, mean (SD) | 2.73 (2.28) | 2.36 (1.34) | 0.49 |
| WOS, mean (SD) | 4.18 (0.40) | 4.29 (0.47) | 0.55 |
| Secondary infection, % present | 2 (18%) | 1(7%) | 0.56 |
| Diabetes Mellitus | 8 (72%) | 6 (43%) | 0.13 |
| Hypertension | 5 (45%) | 6 (43%) | 0.44 |
| CAD | 2 (18%) | 2 (14%) | 0.79 |
| CHF | 3 (43%) | 3 (21%) | 0.73 |
| COPD | 3 (27%) | 0 | 0.03 |

**Figure 1.**
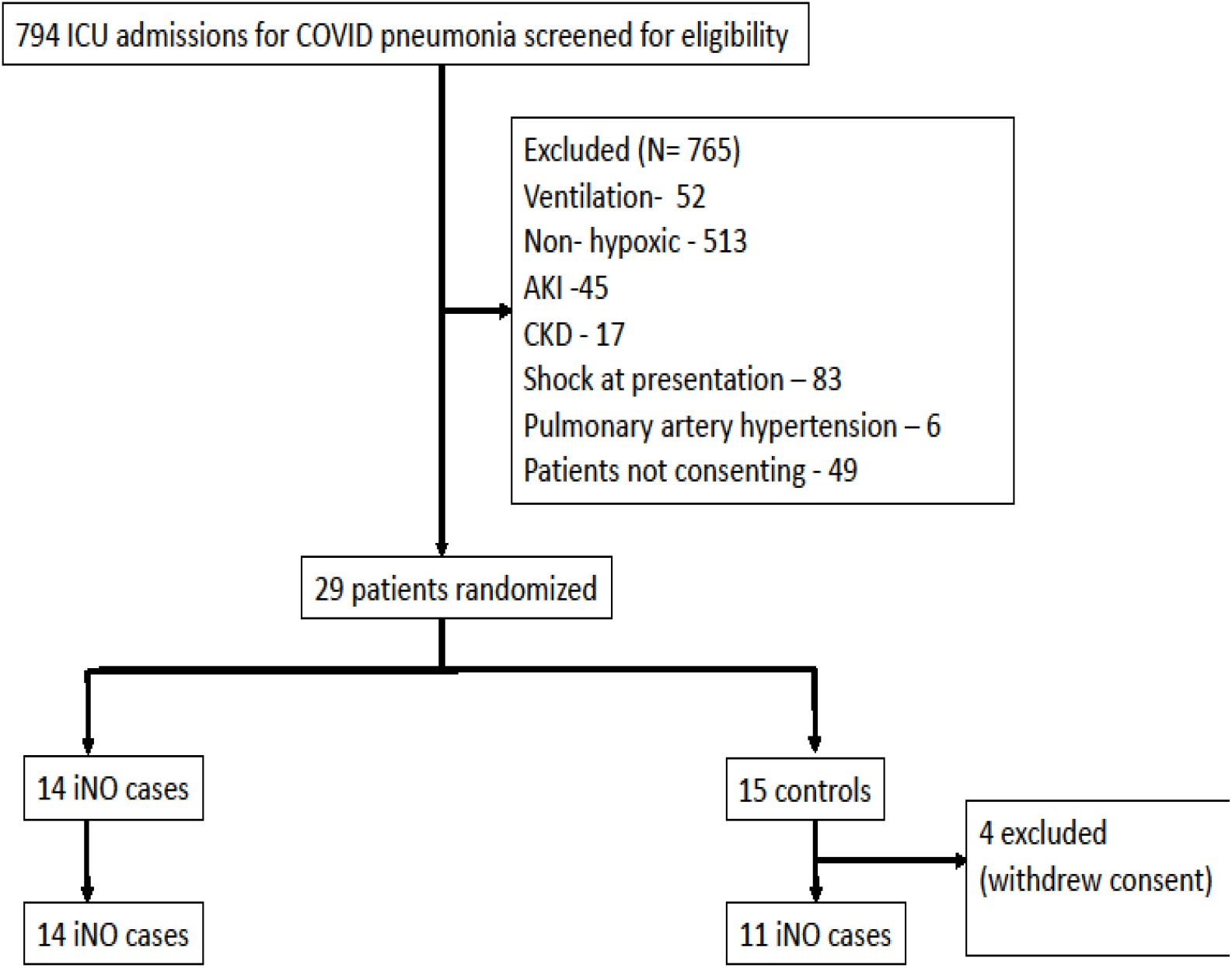
depicts the CONSORT diagram of patient disposition

### Primary outcomes

Viral loads were assessed using nasopharyngeal swabs using quantitative real-time PCR as delineated above. To account for any variability in the sample collection, we plotted normalized Ct values of two genes, ORF1ab and N gene, against an internal control gene Ct value. Of the 25 patients enrolled in the study, 23 were positive for the presence of viral RNA at baseline (Day 0). Longitudinal analysis of viral load at days 0, 3, 5, 7, and 10 shows a significant reduction (p <0.002, N=23) in viral load (inversely correlated to Ct value) in the iNO treated group, as compared to the control group, for both N gene and ORF1ab and this was noted from Day 5 to Day 10 (Figure 2A and 2B). Moreover, the viral load reduction in the iNO treated group was more efficient than in the control group (Table 3). To further evaluate the viral clearance efficiency of iNO, we calculated the time taken by patients in both iNO treated and control groups to reach a Ct value of 35 or above, which corresponds to a mean normalized Ct value of 8.5. A Ct value of above 35 with a mean normalized Ct value of 8.5 is indicative of the patient having clearance of the viral load (36). Notably, 100% of patients in the iNO treated group reached a normalized Ct value of 8.5 or more within five days of iNO treatment, whereas only 72% of patients in the control group cleared the viral load in a comparable time frame (p=0.04) (Figure 3A and 3B). Treatment provided a significant beneficial effect on viral loads during the longitudinal follow up (Table 2).

**Table 2:**
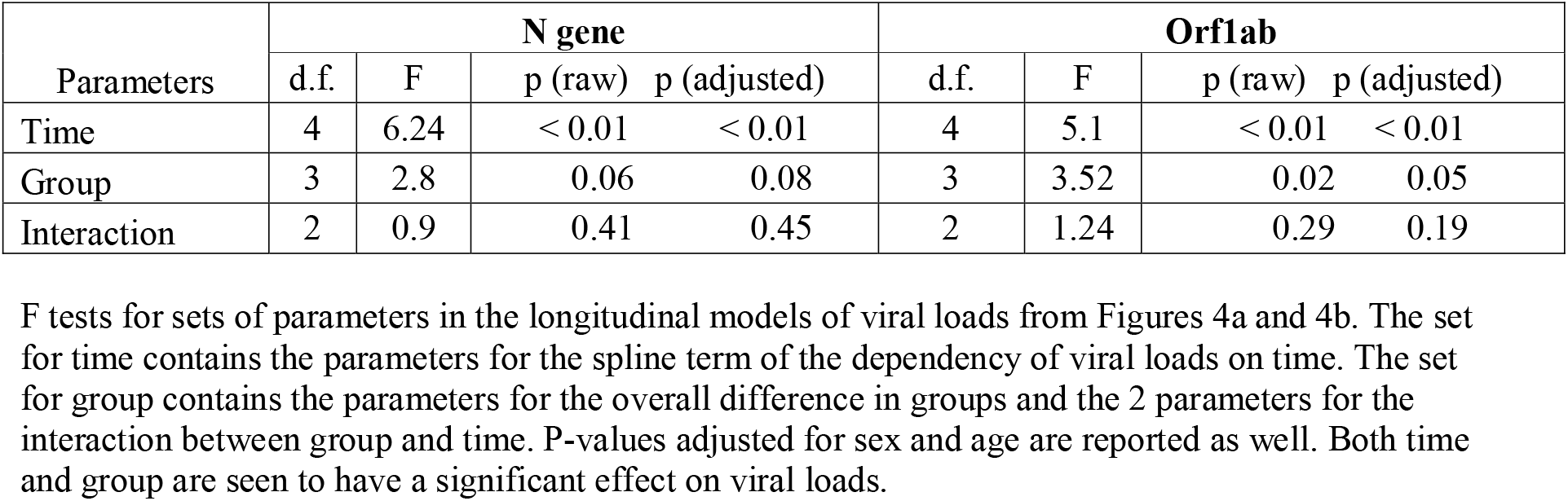
F tests for sets of parameters in the longitudinal models of viral loads from Figures 4a and 4b.

**Table 3:** Primary and secondary outcomes.

| **Outcomes** | **Test**<br>**(N=14)** | **Controls**<br>**(N=11)** | **p** |
| --- | --- | --- | --- |
| Number of patients with 2-point improvement in WOS scale at 7 days | 2(14.2%) | 2(18%) | 0.77 |
| Number of patients with 2-point improvement in WOS scale at 14 days | 11 (78%) | 4(36%) | 0.032 |
| Mean NIV hours | 35.7±54.62 | 10.3±21.8 | 0.06 |
| Need for invasive mechanical ventilation | 0 | 4(36%) | 0.01 |
| Incidence of methaemoglobin levels >3% | 0 | 0 | - |
| 28 day mortality | 0 | 4(36%) | 0.03 |
| Average length of ICU stay | 9.64±3.67 | 11.72±5.29 | 0.12 |
| Average length of hospital stay | 16.21±7.26 | 18.5±9.34 | 0.26 |

**Figure 2A.**
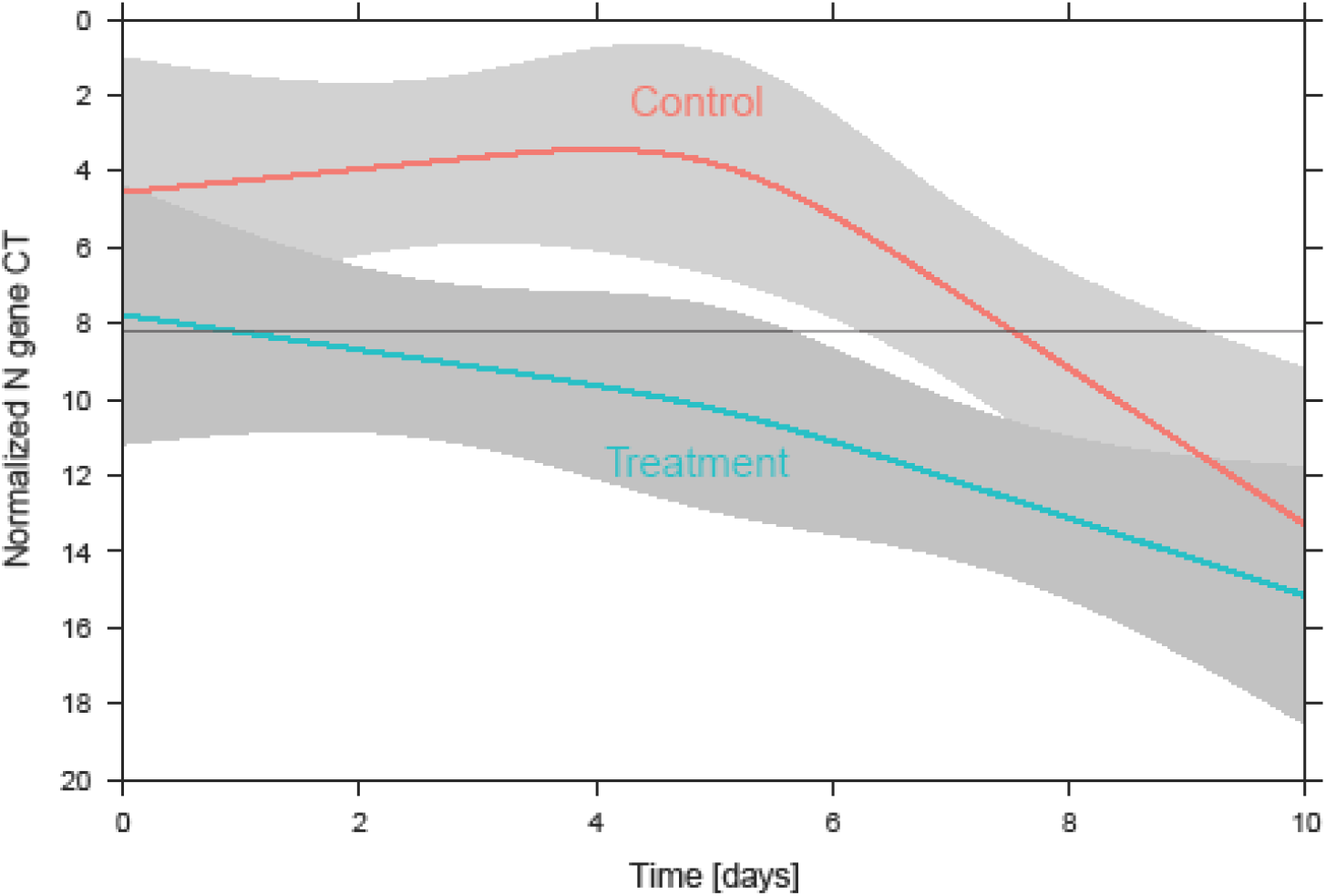
shows the average normalised CT values (N gene) with 95% confidence bands over time for the two groups. The horizontal grey line indicates the CT threshold of 8.205.

**Figure 2B.**
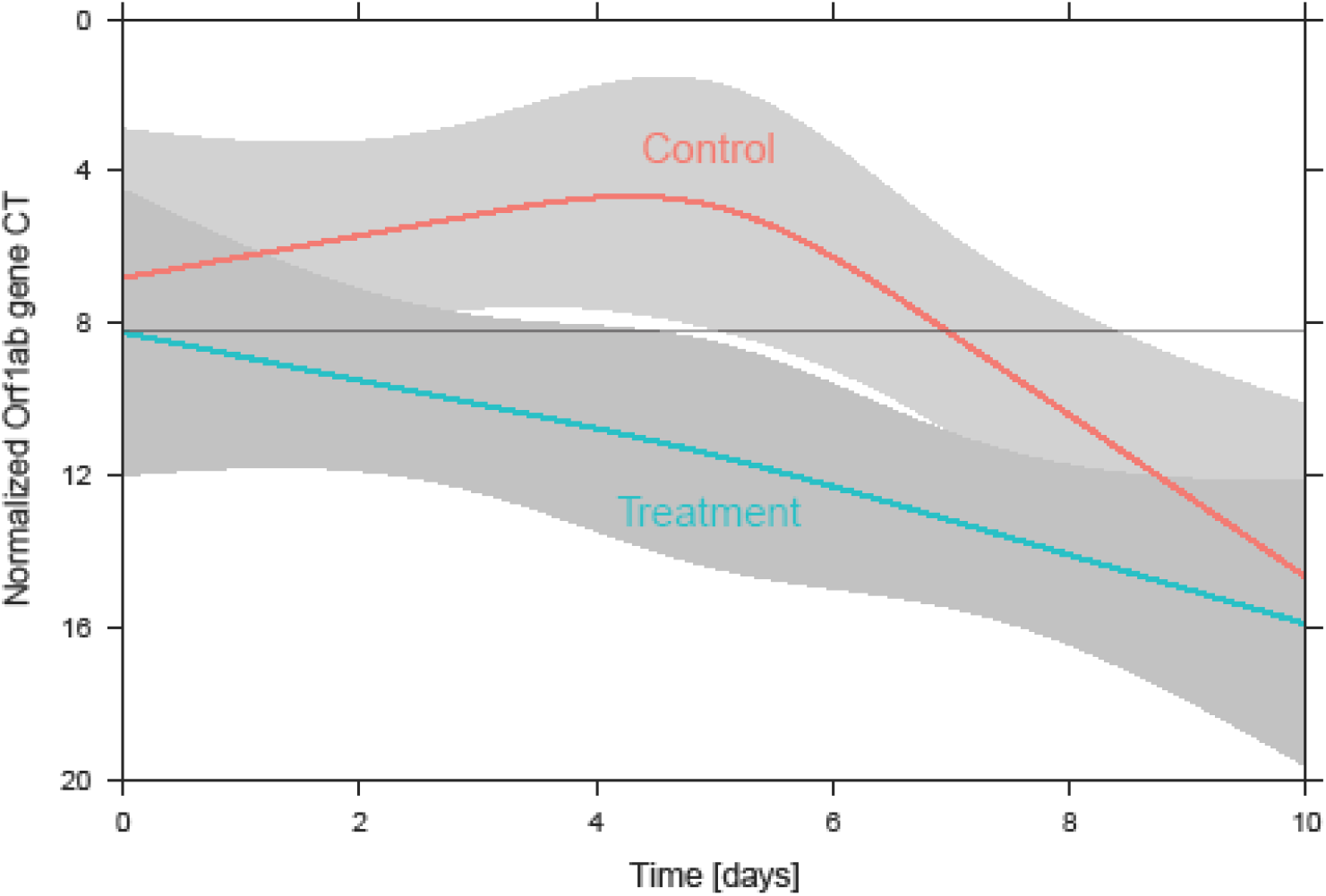
shows the average normalised CT values (Orf1ab gene) with 95% confidence bands over time for the two groups. The horizontal grey line indicates the CT threshold of 8.205.

**Figure 3A:**
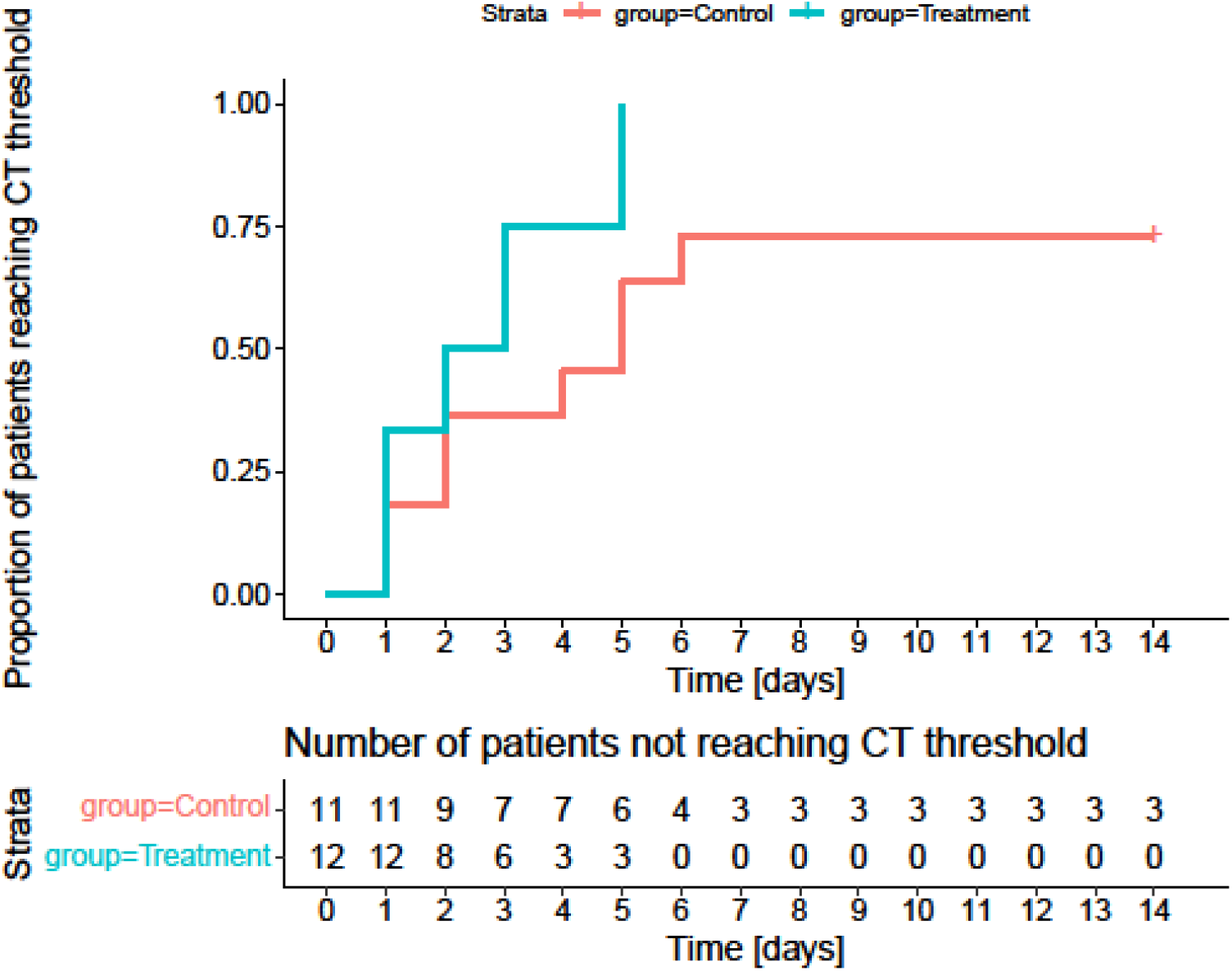
Time till patients reached at least a delta CT value (N gene) of at least 8.205. The proportion of patients that reached a delta CT value (N gene) of at least 8.205 (which corresponds to a CT value of 35) over time (p-values 0.04 for N gene) is depicted.

**Figure 3B:**
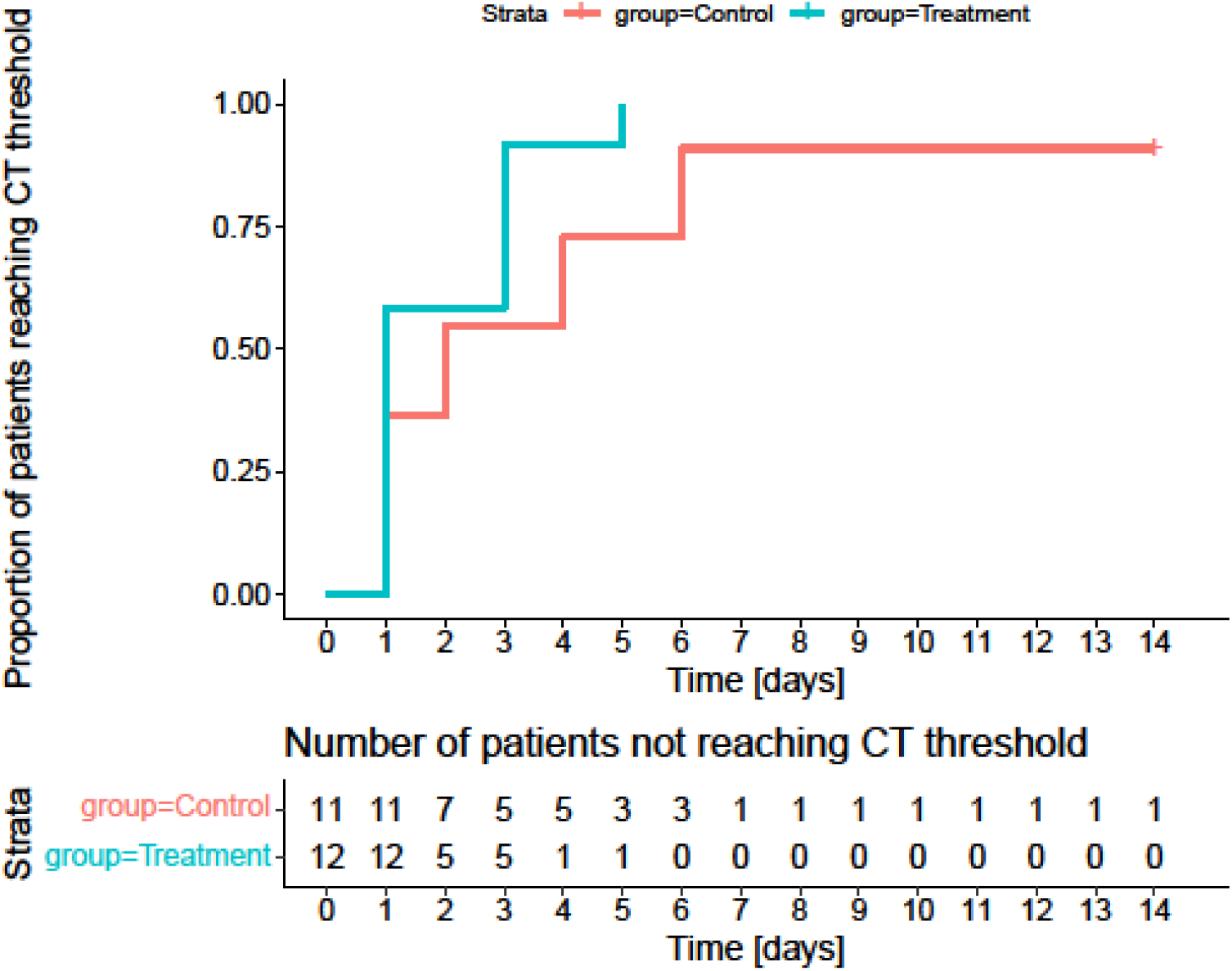
Proportion of patients that reached a delta CT value (Orf1ab gene) of at least 8.205 (which corresponds to a CT value of 35) over time (p-values 0.04 for N gene).

Consistent with this significant treatment benefit, almost all of the patients (13 out of 14; 93%) in the treatment group demonstrated an improvement ≥ 2 points on the WHO Ordinal Scale (WOS), for Severe Acute Respiratory Infections. Of these, 78.5% of patients (11 out of 14), in the iNO treatment group showed an improvement in the WOS from 4 to 0, over the treatment period of 14 days, while the remaining patients showed moderate to mild improvement. However, only 36% (4 out of 11), of the patients in the untreated control group showed an improvement from 4 to 0 in the WOS. The contrast in improvement as adjudged by this parameter was sharpened by the deaths in the control group (n=4, 36%), during the study period (Figure 5A). The median time till a 2 point improvement in WOS was 14 days in the control group and 11 days in the treatment group. The cumulative event, when plotted, also demonstrates a significant 2 point improvement in WOS in the iNO treated patient cohort, as compared to the control (Figure 5B). The differences between the groups persist, even when adjusted for age and sex (Table 4)

**Table 4 :** Estimated daily additional improvement in the treatment group with respect to the WHO ordinal scale.

|  | **Estimate** | **95%**<br>**CI** | **p-**<br>**value** |
| --- | --- | --- | --- |
| Unadjusted | 0.1 | 0.04-0.15 | < 0.01 |
| Adjusted for age and sex | 0.11 | 0.06-0.17 | < 0.01 |
| Adjusted for age, sex,<br>CCI | 0.09 | 0.03-0.16 | < 0.01 |

The estimates are the fixed effects from mixed-effects regression models for the average daily additional improvement due to treatment. The estimates are shown together with 95% confidence intervals and p-values for the hypothesis the treatment and control group have identical improvements.

### Secondary outcomes

A total of 4 patients died in the control group (4 out of 11, 36%) as against no deaths in the treatment group, within the study period (Figure 4). None of the 14 patients in the treatment group were assigned to invasive ventilation during the trial, while in the control group 4 out of 11 patients (36%) received invasive ventilation (p = 0.03). In the treatment group 50% of patients were assigned to non-invasive ventilation, while in the control group the percentage was 36% (p = 0.69). Mean SOFA at 72 hours for the treatment and control group did not reveal a statistically significant difference, while the change in SOFA (ΔSOFA) at 72 hours showed significant improvement in the treatment group (p=0.035). There was no significant difference in the total length of stay in intensive care and in hospital between the two groups (Table 3). None of the patients administered iNO demonstrated levels of methemoglobin >3% during the course of therapy.

**Figure 4:**
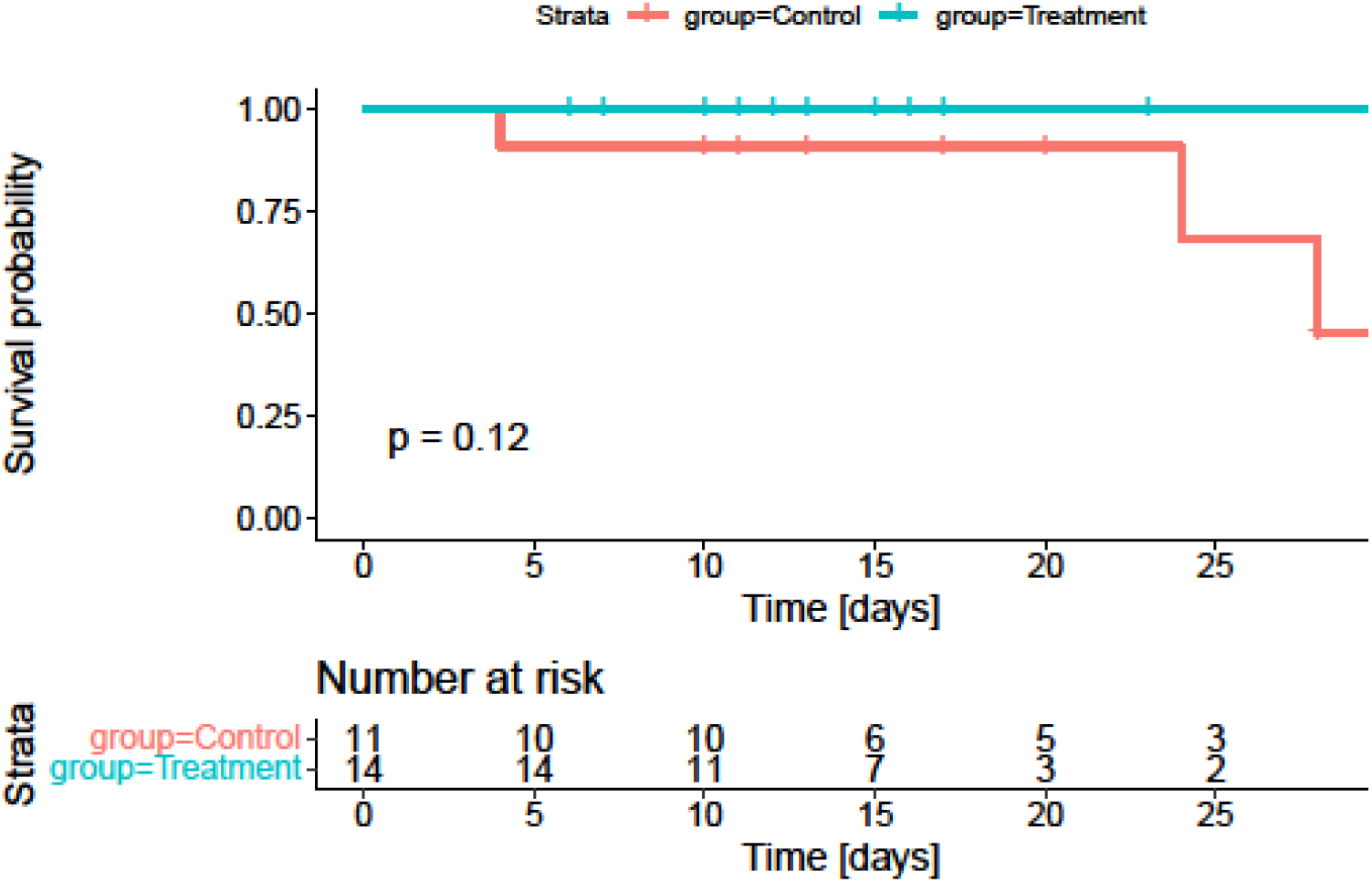
Kaplan-Meier survival plot for iNO treatment and control groups. There was 100% survival of patients in the treatment group during the first 28 days, as compared to 64% survival of patients in the control group (p = 0.03 for difference in the two groups).

**Figure 5A:**
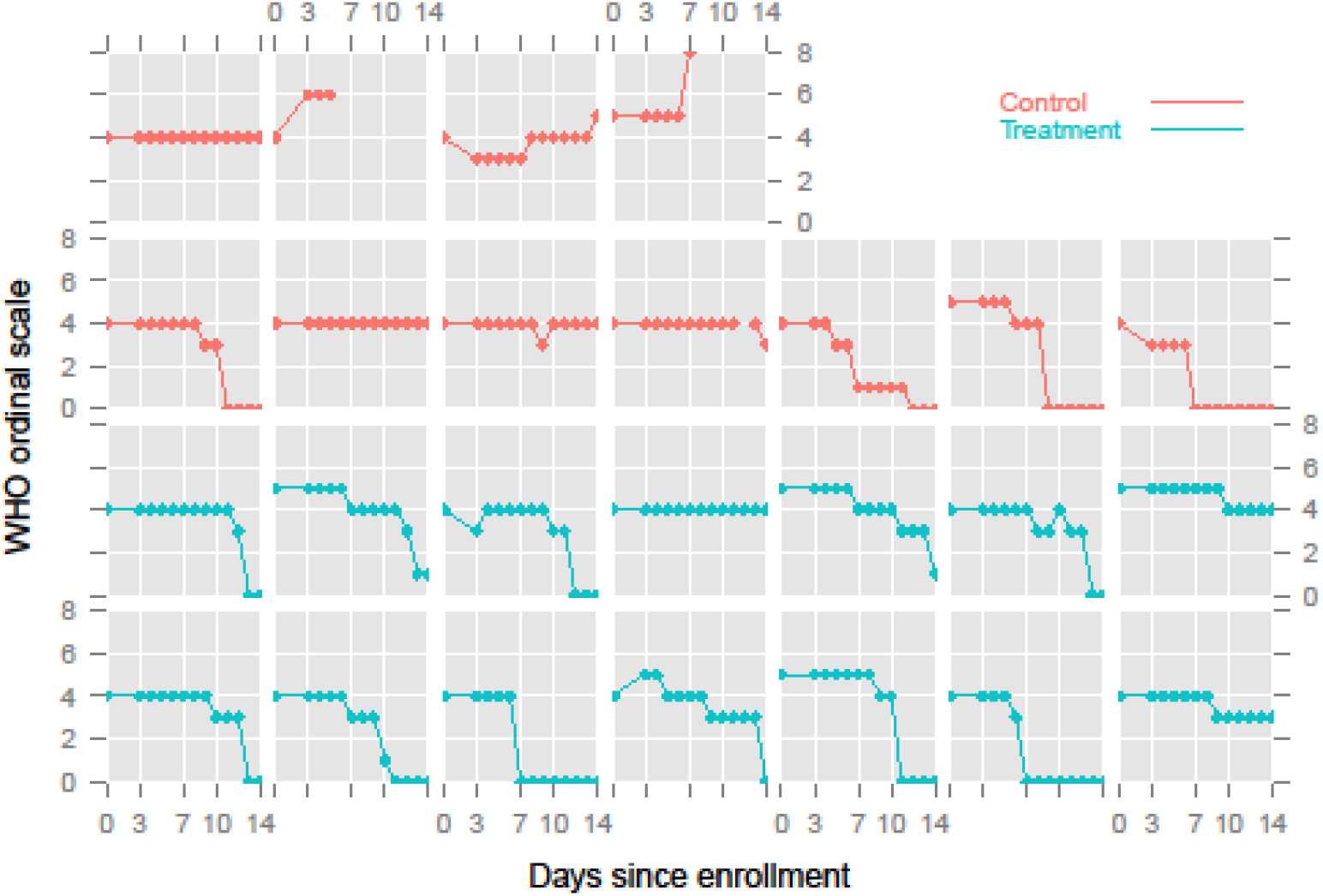
Individual values of the WHO ordinal scales for patients in the treatment group and patients in the control group.

**Figure 5B:**
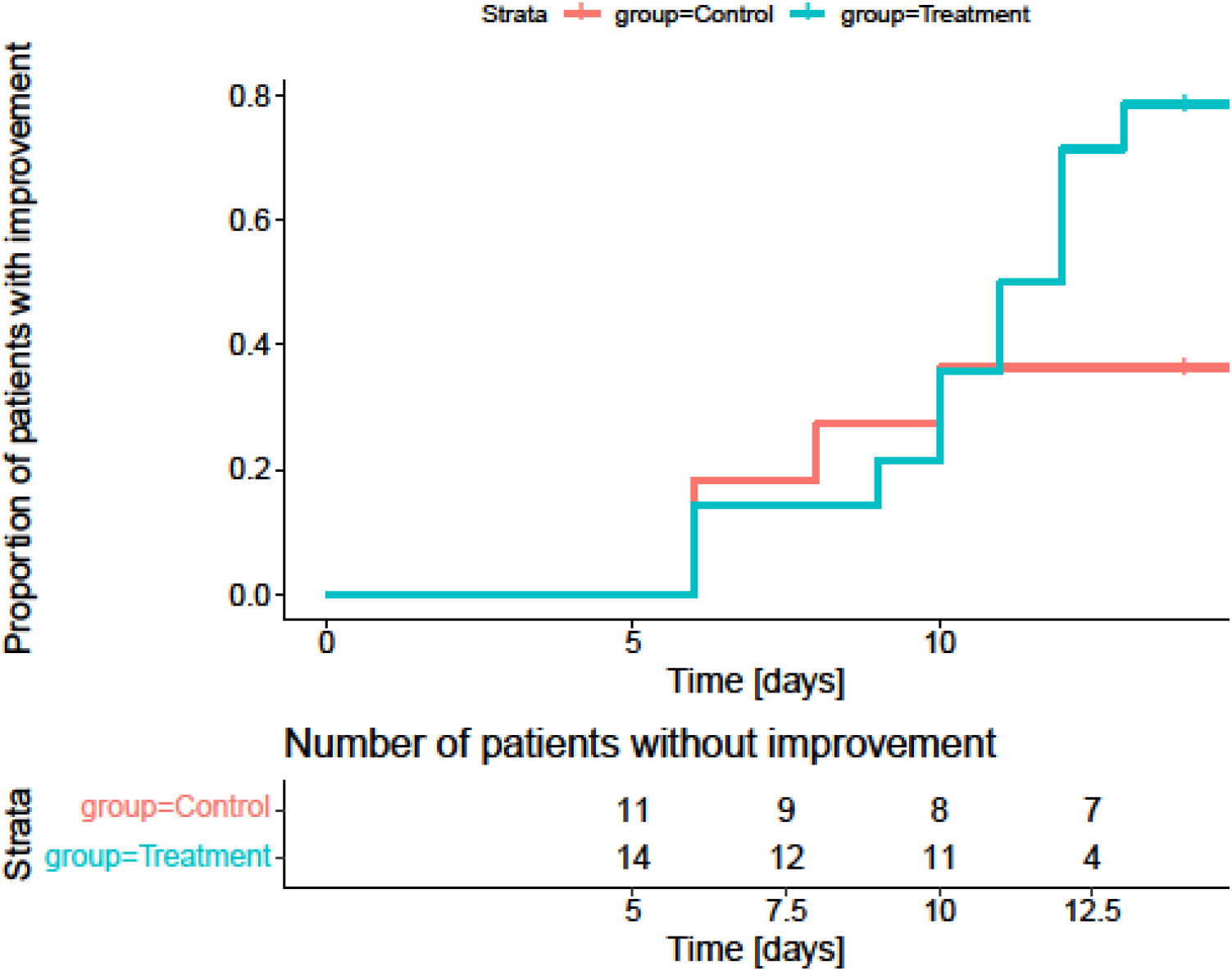
Proportion of patients with an improvement by at least 2 points in the WHO ordinal scale. During the two week study period, in the treatment group, 11 out of 14 (79%) of patients improved by at least 2 units in the WHO ordinal scale, while in the control group 4 out of 11 (36%) patients improved (p = 0.05).

## DISCUSSION

We report here the results of our feasibility trial of inhaled Nitric Oxide (iNO) in the treatment of Covid -19. Although both the treatment and the control groups did receive other antiviral medications, they were equally balanced with respect to these drugs. The study cohort also universally received Dexamethasone as a follow through of the RECOVERY trial (4).

Using the normalised CT ratio for the N gene and the Orf1ab gene as a surrogate of viral load, administration of iNO was associated with significant decline in the viral load. The treatment group showed both more complete and faster efficiency in clearing SARS CoV-2 compared to controls. Nitric oxide is a broad spectrum antimicrobial (15) and NO donors have been shown to inhibit viral RNA replication in a coronavirus model(16), including SARS CoV-2 (17). They inhibit viral entry into host cells via the S protein and its interaction with ACE2 receptor (21). It has also been demonstrated that SARS CoV2 is directly cytopathic to endothelium (32) indicating vasculopathic effects of the virus *in vivo* (33), and the benefits of anticoagulants (34). Nitric oxide occupies a key role in vascular homeostasis with effects on vascular tone and inflammation mediated by endothelial leukocyte interactions (35). (36). The majority of studies on iNO in Covid-19 (20)(21) (22)(23)(24)(25) have focused on its vascular effects; a benefit is also suggested by the higher decline of SOFA scores in the treatment group in this study. However, the use of iNO in improving gas exchange in other models of severe lung injury has been disappointing (37)(38). A single case series has attributed the benefits of iNO in Covid 19 to antiviral effects but offers no proof (26). As such, this report is the first systematic description of the anti-viral properties of iNO.

At the start of our study, a larger albeit insignificant proportion of treatment patients were deemed to require non -invasive ventilatory support. At the conclusion of the study however a significantly larger proportion of the control group progressed to more severe disease, needed invasive mechanical ventilation and died more often. Viewed from the dimension of respiratory symptoms, the treatment group gained faster and more complete improvements on the World Health Organisation 8 point ordinal scale for Severe Acute Respiratory Syndromes. Mechanistically, it has been shown that viral load has clear correlation to the severity of Covid-19 infection (5), to hypoxia (39)and to deaths arising from it (40). We recruited patients with Covid 19 severe enough to warrant oxygen therapy, but not diseased enough to require invasive mechanical ventilation or extracorporeal therapies. We suggest that a key element of antiviral efficacy in our study could have been early institution of inhaled nitric oxide therapy. This hypothesis has support from other virus-induced lung injury models (41) and the secondary analyses of the Adaptive Covid-19 Treatment Trial-1(7)(8)(9).

Despite the high doses of iNO used in this study and by other research groups using iNO for Covid-19 (42)(43), there was no significant incidences of methemoglobinemia, pointing to the safety of pulsed high dose nitric oxide therapy.

This report suffers from several limitations. First, larger conclusions as to the size of the treatment effect cannot be made from this small feasibility trial. Since other antivirals were also used, it cannot be conclusively shown that the antiviral efficacy was predominantly or solely owing to iNO. The doses used in this study use the highest doses available on the SLE Inosys (SLE Surrey, UK) delivery platform and it is uncertain whether a different dosing regimen will be efficacious. Since dexamethasone was also used universally in the study population, clinical improvement may have also accrued to it (4). Finally, it is uncertain whether iNO would be equally efficacious in sicker patients.

In sum, we offer the first composite evidence of the utility of inhaled Nitric Oxide on accelerating viral clearance of SARS CoV-2 infection in hypoxemic patients with Covid-19. Its use appears to improve clinical recovery without any attendant harms and the results of this trial should be extended in a larger trial population.

## Data Availability

The datasets generated during and/or analysed during the current study are available from the corresponding author on reasonable request.

## Author contributors

BN and GK contributed to conceptualisation, formal analysis, funding acquisition, investigation, methodology, project administration, resources, supervision, validation, visualisation, writing – original draft, and writing – review & editing. ICP contributed to conceptualisation, writing – original draft, and writing – review & editing. AJ, MM, DTS, VM and TM contributed to conceptualisation, formal analysis, funding acquisition, investigation, methodology, project administration, resources, supervision, validation, visualisation, writing – original draft, and writing – review & editing. DSK and PP contributed to data curation, formal analysis, project administration, supervision, validation, methodology. GG and FE contributed to formal analysis, software, visualisation, writing – original draft, and writing – review & editing. RJ, SJ and AK contributed to data curation, investigation, validation. Data verified by AJ, DTS and MM.

## Acknowledgements

We acknowledge Dr. Subramania Iyer, Dr Nandita Mishra, Bri Sai Bala Madathil, Dr Zubair Umer Mohamed, Dr Perraju Bendapudi and Dr Shyam Sundar P and Dr Sindhu Balakrishnan for their critical insights into study design and conduct of the study.

## Disclaimer

The funders had no role in study design, data collection and analysis, decision to publish, or preparation of the manuscript.

## Funding sources

The work was supported by the collaborating centre, the Amrita School of Biotechnology, Amrita Vishwa Vidyapeetham.

## Potential conflicts of interest

The authors report no conflicts of interest. All authors have submitted the ICMJE Form for Disclosure of Potential Conflicts of Interest.

## Notes

### Competing Interest Statement

The authors have declared no competing interest.

### Clinical Trial

ISRCTN 16806663

### Author Declarations

Drugs Controller General India Institutional Ethics Committee, Amrita Institute of Medical Science

